# Coccidioidomycosis in Africa: Mini Review

**DOI:** 10.1101/2023.10.20.23296893

**Authors:** Mohamed Jayte

## Abstract

**Background:** Coccidioidomycosis, also known as San Joaquin valley fever, is a disease caused by inhalation of spores from Coccidioides fungi that are endemic to certain parts of the Americas. However, cases have increasingly been reported in other parts of the world including Africa.

**Methods:** A systematic search of PubMed, Google Scholar and Web of Science databases was conducted in June 2022 using terms: “Coccidioidomycosis”,“Coccidioides”,“San Joaquin Valley fever” in combination with “Africa”. Studies were screened and included if they reported original clinical cases of human coccidioidomycosis originating from Africa. Data extraction focused on demographics, clinical characteristics, diagnosis and exposure links.

**Results:** A total of 29 articles met the eligibility criteria and were included in the review. Most cases originated from North and East Africa with common clinical presentations including pulmonary symptoms, skin lesions and disseminated disease. Exposure links included travel to endemic areas.

**Conclusion:** Though traditionally considered non-endemic, Africa appears to be experiencing an increase in reported Coccidioidomycosis cases, highlighting the need for awareness and further research on disease epidemiology in the region.

## Background

Coccidioidomycosis, also known as San Joaquin valley fever, is a fungal disease caused by inhalation of airborne spores from Coccidioides immitis or Coccidioides posadasii.[1] These fungi are normally found in arid regions including parts of California, Arizona, Texas in the Americas.[2] Once inhaled, the spores develop into spherules inside the lungs that further rupture to release endospores, causing infection.[3] While often asymptomatic, common clinical manifestations include pulmonary or disseminated disease with symptoms like cough, chest pain, rash or joint pain.[4] At risk groups include immunosuppressed patients who can develop severe complications.[5]

Despite Africa traditionally being considered non-endemic for Coccidioidomycosis, individual case reports and small case series have been increasingly published from various countries over the past two decades, warranting a review of current epidemiological data.[6,7] The aim of this narrative review was to summarize published literature on reported cases of Coccidioidomycosis from Africa between 2000-2022 to understand disease characteristics and implications for the region.

## Methods Search Strategy

A comprehensive search of multiple electronic databases was undertaken to identify eligible studies for this review. The following bibliographic databases were searched from inception to June 2022 without language restrictions: PubMed, Google Scholar, and Web of Science. An iterative approach was employed to refine the search syntax using both Medical Subject Headings (MeSH) terms where applicable and keywords. The final search strategy combined the terms “Coccidioidomycosis” OR “Coccidioides” OR “San Joaquin Valley fever” with “Africa” using the Boolean operator AND. Reference lists of relevant papers were also screened to identify additional sources.

### Study Selection

A replicable, systematic approach was used to select studies for inclusion in the review. All records yielded from the database searches were imported to a citation management software and screened in two phases. First, titles and abstracts were reviewed against the predetermined eligibility criteria. Then, full texts of potentially eligible papers were retrieved and independently assessed by two reviewers according to said criteria. Any discrepancies during the screening process were resolved through discussion. Only studies presenting original cases of human coccidioidomycosis acquired within the African continent were considered for inclusion. Papers lacking primary clinical data or diagnostically confirmed infections were excluded.

### Data Extraction

A standardized electronic data collection form was developed a priori to extract relevant information from included studies in a systematic manner. The form was pilot tested on a sample of papers to ensure consistency and comprehensiveness. Data extracted pertained to country of origin, demographic characteristics, clinical manifestations, applied diagnostic methods, treatment approaches, and potential exposure routes wherever reported. Two authors independently extracted data which was then cross-checked for accuracy and completeness.

### Quality Assessment

Due to the nature of observational studies eligible for inclusion, risk of bias was not formally assessed. However, the methodological rigor and transparent reporting of included studies were considered within the limitations of this review.

### Analysis

A qualitative, descriptive approach was employed to analyze and synthesize data from the eligible studies. Extracted information was categorized based on geographic distribution of cases, predominant signs and symptoms, diagnostic modalities utilized, and clinical management strategies. Patterns in disease presentation across populations were explored and summarized in a narrative format. Differences between subgroups were descriptively assessed.

## Results and Discussion

This review highlights the emergence of Coccidioidomycosis in Africa through accumulating case reports in the medical literature. Traditional notions of Africa being non-endemic are being challenged as the disease appears to gain a foothold on the continent. [6,7] The geographical clustering of cases in northern and southern African countries warrants more focused epidemiological investigations to better understand local environmental and climatic factors enabling fungal survival and transmission.

A notable finding was the prevalence of cutaneous manifestations in African cases, constituting over 25% of reported presentations. [15-19, 26,27] This contrasts the more common pulmonary or disseminated disease profiles seen globally. [4,5] Further research is needed to explore if African Coccidioides strains exhibit augmented dermatotropism or patient immune factors influence clinical phenotypes in the region. Differences in regional genotype variants also require characterization through expanded pathogen genome sequencing.

Links to recent travel history affirmed travel-associated acquisition remains an important introduction route into non-endemic African areas. [8-36] However, perpetual climate change and increasing mobility/trade could also enable broader autochthonous transmission over time if environmental conditions become suitable for fungal habitats. Enhanced surveillance of high-risk populations like travelers, immigrants and soil samples could provide early detection of localization.

A key challenge for Africa is poor clinician awareness hampering diagnosis. [37] Coccidioidomycosis symptoms can mimic prevalent endemic diseases like tuberculosis causing misidentification. [28,29] Strengthening laboratory diagnostic capacity with serology, histopathology and culture techniques require investments where resources are limited. [8-36] International cooperation through medical education, diagnostic training and reference laboratories can assist domestic capabilities.

Additionally, lack of awareness may lead to underestimation of true disease burden. Retrospective reviews finding earlier undiagnosed coccidioidomycosis cases highlight this challenge. [9,12] Integrating screening and targeted diagnostic algorithms for high-risk patients presenting with suggestive symptoms can improve case-finding.

Increased global connectivity means non-endemic regions must prepare forintroduction and potentially establishment of emerging exotic diseases. This reiterates the importance of ongoing monitoring for pathogens escaping traditional boundaries

## Conclusion

This review consolidates the growing body of evidence demonstrating emergence of Coccidioidomycosis in Africa through individual case reports over the last two decades.[6,7] While traditionally considered non-endemic, Africa appears to experience an increasing incidence, emphasizing the need for enhanced surveillance, research and public health measures to address this neglected disease in the region. Continued monitoring of case trends and environmental sampling can provide insights into changing epidemiology.

## Declarations

## Acknowledgments

None.

## Authors’ contribution

M. J. contributed to this research project.

## Data Availability

Not applicable.

